# Effects of diacutaneous fibrolysis in patients with tension-type headache: A randomized controlled trial

**DOI:** 10.1101/2022.08.25.22278916

**Authors:** Sara Cabanillas-Barea, Silvia Pérez-Guillén, Carlos López-de-Celis, Jacobo Rodríguez-Sanz, Pablo Fanlo-Mazas, Andoni Carrasco-Uribarren

**Author notes:** Corresponding author: (SPG).

## Abstract

**Background:** Manual therapy appears to be effective for the relief of tension-type headache (TTH), just as diacutaneous fibrolysis (DF) has shown to be a beneficial technique for the relief of symptoms in other dysfunctions. However, no studies have evaluated the potential beneficial effect of DF in TTH. The aim of this study is to analyze the effect of three sessions of DF in patients with TTH.

**Methods:** Randomized controlled trial in 86 subjects (43 intervention/ 43 control group). The headache frequency, the headache intensity, the pressure pain thresholds (PPTs) at trapeziometacarpal joint, upper trapezius, suboccipital, frontal and temporal muscles, parietal sutures and the cervical mobility were measured at baseline, at the end of the third intervention and one-month after the last intervention.

**Results:** Statistically significant differences with p values <0.05 were observed between groups in favor of the intervention group in the one-month follow-up in the following variables: headache frequency, headache intensity, flexion, extension, right and left side-bending, right and left rotation, PPTs in left trapeziometacarpal joint, right suboccipital muscle, right and left temporal muscle, left frontal muscle and right and left parietal.

**Conclusions:** DF provides a beneficial effect in reducing headache frequency, relieving pain, and improving cervical mobility in patients with TTH.

## Introduction

Tension-type headache is the most common primary headache worldwide [1] and equals migraine in cost [2]. Symptoms can include bilateral pain in frontal and occipital regions, dull pain across the forehead, sides or back of the head and tenderness on the scalp or muscles of the neck, upper back, shoulders, and jaw [3,4].

The most common abnormal finding in these patients is the increased sensitivity of the pericranial structures; this sensitivity seems to increase uniformly throughout the cranial and cervical region, affecting skin, muscle, tendon, and fascial tissue [5]. In addition to the characteristics of pain, decreased cervical range of motion, decreased strength of the cervical muscles and excessive forward head position have been found [6].

Therapeutic approaches include pharmacological and non-pharmacological interventions [7]. For many individuals with tension-type headache, pharmacotherapy’s mainstays are simple analgesics and nonsteroidal inflammatory drugs [8]. Despite this, the European Federation of Neurological Societies guideline promotes non-pharmacologic therapies, having fewer side effects than pharmacological treatments [9]. Non-pharmacologic therapies may be a valid therapeutic option for subjects with headaches, despite their limited scientific basis [10], and the lack of evidence on the effectiveness of physical therapy options [11].

Physical therapy is the most used non-pharmacological treatment of tension-type headache and includes several treatment strategies such as manual therapy, postural exercise, relaxation, physical training, and mixed physiotherapy approaches [2,12]. Manual therapies appear to be the most common non-medical treatment used to manage common recurrent headaches [4]. Within manual therapy techniques, soft tissue treatment resulted in a significant and beneficial effect on the pain intensity and frequency in these patients [12,13].

Diacutaneous fibrolysis is a physiotherapeutic technique developed following Cyriax Deep Friction Massage principles. A set of metallic hooks is used to achieve a more in-depth and more precise application than manually. A recent systematic review and meta-analysis published by Cadellans et al. [14] has shown that diacutaneous fibrolysis is an effective technique for improving symptoms and function in other musculoskeletal disorders such as shoulder pain [15,16], chronic lateral epicondylalgia [17], patellofemoral pain [18] and carpal tunnel syndrome [19,20].

The body of evidence supporting the clinical effectiveness [16–18,20] of diacutaneous fibrolysis has increased in recent years. Nevertheless, to the best of the authors’ knowledge, no study to date has investigated the effects of diacutaneous fibrolysis in patients with tension-type headache. Therefore, the purpose of this randomized controlled trial was to evaluate the effects of diacutaneous fibrolysis on headache frequency, intensity, cervical pressure pain thresholds (PPTs) and cervical mobility in patients with tension-type headache. The hypothesis is that three sessions of diacutaneous fibrolysis would effectively reduce headache frequency and intensity, tissue sensitivity, and improve cervical range of motion in tension-type headache patients.

## Materials and methods

### Study Design

The study design was a 2-group randomized controlled trial with pre-, post-intervention and one-month follow-up measurements. The allocation ratio was 1:1. The study was performed at University of Zaragoza from October 2015 to May 2018. A researcher not involved in the study randomized the intervention groups using a computer software (www.random.org). The results were placed in a concealed opaque envelope and were assigned to participants. The study was conducted following the Declaration of Helsinki and the approval of the Clinical Research Ethics Committee of the Community of Aragon (CEICA), Spain (CEICA number: PI15/229) was obtained. All participants provided informed consent before their enrolment in the study. This clinical trial was carried out in the Faculty of Health Sciences facilities, University of Zaragoza, Spain (clinicaltrials.gov number: NCT03056131). This study followed the CONSORT criteria.

### Simple sizes calculation

The sample size was calculated based on the frequency of days with headache outcome, obtaining the highest number of subjects (43 subjects per group), making a total sample of at least 86 subjects. The sample size was calculated using the GRANMO 7.12 program with a α risk of 0.05, two-sided test and a β risk of 0.20. For frequency of headache, an estimated common standard deviation of 4.6 and a minimum expected difference of 2.8 were used [21].

### Subjects

Ninety-three subjects diagnosed with TTH were recruited form primary care. The inclusion criteria were age>18 and a diagnosis of tension-type headache in adherence to the International Classification of Headache Disorders (ICHD) criteria: headache occurring on ≥15 days/month on average for >3 months, headache lasting from 30 minutes to 7 days, bilateral location, pressing/tightening quality, mild or moderate pain, not aggravated by routine physical activity, no nausea or vomiting and no more than one photophobia or phonophobia [22]. Participants were excluded if they had damaged skin, cutaneous lesions or vascular abnormalities in the cranio-cervical area, concomitant treatment with platelet antiaggregant agents, previous cervical or cranial surgery and patients with pending litigation or court claim.

### Procedure

After baseline examination, participants were randomly allocated to the control (n=43) or intervention (n=43) group using a computer-generated sequence of numbers (simple randomization) using Microsoft Excel performed by an independent blinded investigator.

Allocation was made by an assistant who was not informed about the random sequence. Participants received a sealed and numbered envelope. This assistant then scheduled the participants for the first treatment session.

The total duration of the study was 6 weeks, with four face-to-face sessions. The first one was provided to record information from participants and to measure the outcomes for this study after the inclusion at baseline (T0). The post-treatment assessment was per-formed after the final session of intervention protocol (T1) and the follow-up assessment was performed after one month (T2). The assessment was carried out by the same person – a trained physiotherapist – throughout the study, following a standardized procedure. The evaluator was kept uninformed about the subjects’ headache history.

### Measurements

The primary outcome was the frequency of the episodes of headache per month. As secondary outcomes the headache intensity, the pressure pain thresholds and cervical mobility were measured.

For the frequency of headache episodes, the headache days suffered during the last month were recorded. This variable was recorded at times T0 and T2.

The headache intensity was registered with a visual analogue scale, which is a valid and reliable tool for measuring headache pain intensity [23]. A 10 cm vertical line was used, with a descriptor at each end of the line (“no pain” and “worst pain imaginable”). The headache intensity was recorded in four different instants: actual pain, usual pain, worse and better in the last month.

Pressure pain threshold was measured using a handheld pressure algometer (Somedic AB, Farsta, Sweden) at different cervical region sites to reflect localized hyperalgesia and on the hand to reflect distal hyperalgesia. The probe (1 cm2) was placed right angle to the skin. The pressure was applied at a rate of 30 KPa/s, and participants were instructed to indicate when the sensation changed from a sensation of pressure to the first sensation of pain. The subject was in a supine position, and the pressure pain thresholds were assessed in trapeziometacarpal joint, suboccipital muscles, frontal muscles, temporal muscles, and parietal suture. All sites were first located and marked. The test sites on the neck and head were located based on bony landmarks. Patients press the button of the digital algometer at the precise moment that pressure sensation changed to pain. The reliability of this measurement is high (ICC = 0.91-0.97) [24].

The CROM (floating compass; Plastimo Airguide, Inc, Buffalo Groove, IL) device was used to measure the range of movement of the cervical spine, which has shown to have good intra- and interexaminer reliability (ICC > 0.80) in all the measurements [25]. The general active range of motion of the cervical spine was assessed in a sitting position with the back resting on the chair’s backrest. Flexion, extension, left and right side-bending and left and right rotation were registered. The flexion and extension of the upper cervical spine (UCS) were measured to stand with the head resting on a wall [26].

The Global Rating of Change Scale (GROC-Scale) was rated according to the question: “Compared to prior inclusion in the study, how much has your headache problem changed?” (Seven for improvement, seven for deterioration and one for without changes). The minimal clinically significant difference in patients with headache is ±4 on the GROC-scale. After the registration, the results were grouped into three categories: without clinical change, with clinical worsening and with clinical improvement [27].

All these variables were recorded in T0, T1 and T2, except for the frequency, the visual analogue scale “worst moment”, “best moment” and “usual pain” that were registered in T0 and T2. The GROC-scale was assessed at T1 and T2 follow-ups.

### Intervention

The intervention had a 1-week duration, with three face-to-face sessions of diacutaneous fibrolysis. The intervention was about 30-minute-long and was applied with at least 1-day separation between sessions. The physiotherapist who applied the intervention had more than ten years of experience in using the technique.

The diacutaneous fibrolysis was performed in the intermuscular septum between muscles with an anatomical or functional relationship with the cervical spine. Those muscles were trapezius, levator scapulae, splenius cervicis, splenius capitis, and sternocleidomastoid. The technique was also applied on the bony edges of the dorsal spinous processes, the scapula, and the base of the skull. The diacutaneous fibrolysis intervention was applied in a centripetal direction towards the head (pain localization), starting at a distance from the thoracic region, continuing through the scapular region, and finally ending in the cervical and cranial region. The diacutaneous fibrolysis treatment involved three main techniques: treating painful points by pressure inhibition, separating different muscle planes, and scratching tendon attachments (Fig 1). The patient was changing position (supine, lateral and prone position) depending on the area to be treated.

**Fig 1.** Diacutaneous fibrolysis techniques. 1. Treating painful points by pressure inhibition, 2 and 3. Separating different muscle planes and 4. Scratching tendon attachments.

The control group remained in the supine position for 15 minutes without receiving intervention. The placebo effect has already been studied with the simulated diacutaneous fibrolysis technique [15]. A sham technique was avoided as in other studies that have applied sham diacutaneous fibrolysis, when participants experienced a worsening of their symptoms [20].

### Statistical Analysis

Statistical analysis was conducted with the SPSS 20.0 package (IBM, Armonk, New York). The mean and standard deviation were calculated for each variable. The Kolmogorov-Smirnov test was used to determine the normal distribution of quantitative data and the Chi-Square test for sex variable (p>0.05). Within-group and between-group differences were analyzed using the Student t-test and for the quantitative data Fisher test. Besides, the effect sizes were calculated using Cohen’s d coefficient. An effect size >0.8 was considered large; about 0.5, intermediate; and <0.2, small. A p value <0.05 was considered significant.

## Results

Ninety-three subjects were recruited between October 2015 to May 2018. Eighty-six met the selection criteria. Five subjects (four did not complete the intervention protocol and one did not attend the evaluation session) dropped out the study, so the data analysis was completed with seventy-nine participants (57 females and 22 males, 38.35 age ±15.78). Subjects were randomly allocated to one group or another (Fig 2).

**Fig 2.** Flowchart of patient recruitment.

The demographic characteristics are summarized in Table 1. No subject reported adverse effects during or at the end of the intervention.

**Table 1.**
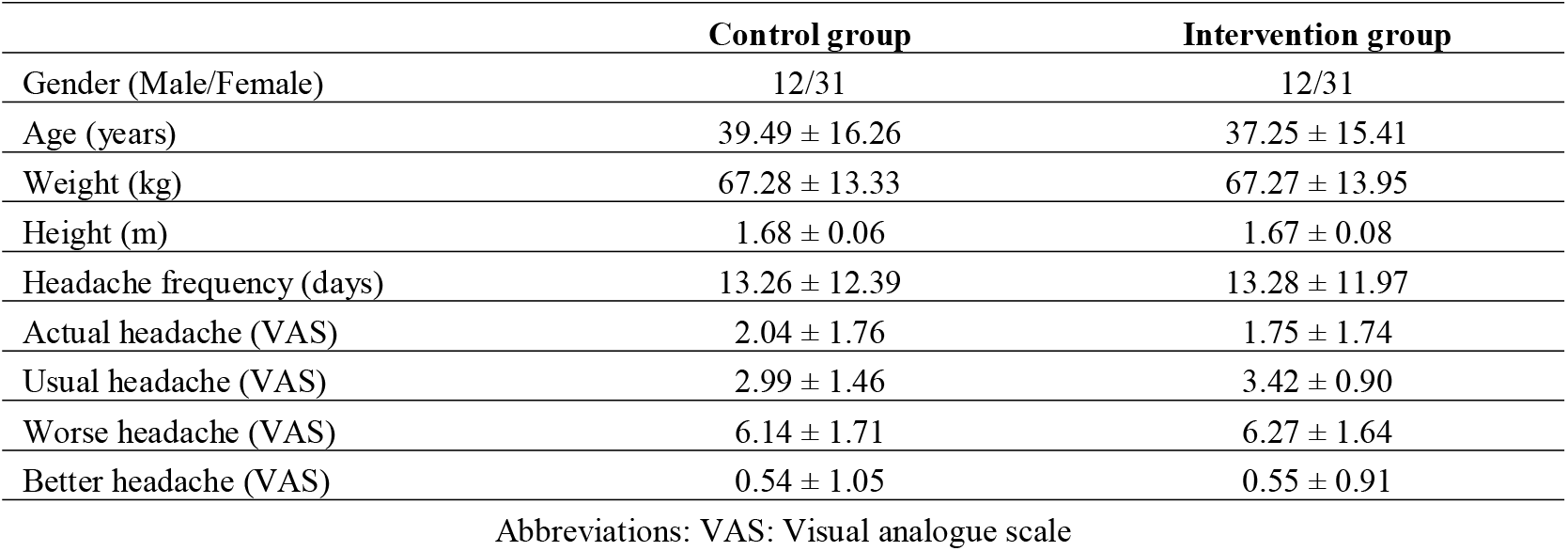
Subject demographic characteristics at baseline

|  | Control group | Intervention group |
| --- | --- | --- |
| Gender (Male/Female) | 12/31 | 12/31 |
| Age (years) | 39.49 ± 16.26 | 37.25 ± 15.41 |
| Weight (kg) | 67.28 ± 13.33 | 67.27 ± 13.95 |
| Height (m) | 1.68 ± 0.06 | 1.67 ± 0.08 |
| Headache frequency (days) | 13.26 ± 12.39 | 13.28 ± 11.97 |
| Actual headache (VAS) | 2.04 ± 1.76 | 1.75 ± 1.74 |
| Usual headache (VAS) | 2.99 ± 1.46 | 3.42 ± 0.90 |
| Worse headache (VAS) | 6.14 ± 1.71 | 6.27 ± 1.64 |
| Better headache (VAS) | 0.54 ± 1.05 | 0.55 ± 0.91 |

Abbreviations: VAS: Visual analogue scale

### Frequency of headache

Headache frequency was significantly reduced in the intervention group (p<0,001) and a change was observed in the control group (p=0,039) but showing an increase.

### Headache intensity

Subjects in the intervention group reported a lower intensity in T1 and T2 follow-ups in actual headache pain. In both cases, statistically significant differences were obtained in T1 follow-up p<0.002 and intermediate effect (d=0.67) and T2 p<0.009. Differences were also found between both groups at T1, p<0.001 and T2 p<0.001 with a large size effect at T1 (d=0.83).

The usual and worst headache pain showed a statistical improvement in the intervention group in T2 p<0.001. The better headache pain showed a statistically significant decrease in the intervention group p<0.010 at T2. No difference was observed in the control group. The usual pain between groups showed a p<0.001 and a large size effect (d=1.56) favoring the intervention group.

### Cervical ROM

The intervention group obtained an improvement in all directions. At T1 follow-up flexion (p<0.017; d=0.44), extension (p<0.001; d=0.67), right side-bending (p<0.001; d=0.66), left side-bending (p<0.001; d=0.80), right rotation (p<0.001; d=0.87), left rotation (p<0.001; d=0.78), UCS flexion (p<0.001; d=0.64) and UCS extension (p<0.001; d=0.56). The control group obtained a statistically significant worsening at T1 in flexion (p<0.001; d=0.55) and extension (p<0.023; d=0.43).

Between group analysis showed difference in favor of the intervention group at T2 follow up in flexion (p<0.001; d=0.74), extension (p<0.001; d=1.06), right side-bending (p<0.001; d=0.77), left side-bending (p<0.001; d=0.69), right rotation (p<0.001; d=0.82), left rotation (p<0.001; d=0.92), UCS flexion (p<0.001; d=0.50) and UCS extension (p<0.003; d=0.45).

#### 3.4. Pressure Pain Threshold

Between group analysis showed difference in favor of the intervention group at T1 follow up in left trapeziometacarpal joint (p=0.003; d=0.41), right suboccipital muscle (p=0.042; d=0.33), right (p=0.003; d=0.57) and left (p=0.011; d=0.49) temporal muscle. At T2 follow up in left trapeziometacarpal joint (p=0.001; d=0.69), right suboccipital muscle (p=0.008; d=0.49), right (p=0.022; d=0.52) and left temporal muscle (p=0.005; d=0.71), left frontal muscle (p=0.008; d=0.52) and right (p=0.012; d=0.47) and left (p=0.050; d=0.42) parietal suture.

### Global rating of change scale

The GROC-scale showed a clinical improvement in 77.5% of the subjects in the intervention group and 2.6% of the participants in the control group at T1 follow-up (p<0.001). In T2 follow-up, 77.5% of the participants felt clinically better in the intervention group nd 5.1% in the control group (p<0.001). Table 2 show the comparison of changes and effect sizes over time for each treatment group.

**Table 2.** Comparison of changes and effect sizes over time for each treatment group.

|  | Baseline<br>Mean ± SD | Follow up T1<br>Mean ± SD | Difference between Baseline |  |  | Follow up T2<br>Mean ± SD | Difference Between Baseline |  |  |
| --- | --- | --- | --- | --- | --- | --- | --- | --- | --- |
|  |  |  | Mean (95% CI) | p<sup>a</sup> | ES |  | Mean (95% CI) | p<sup>b</sup> | ES |
| Frequency of H.<br>Control<br>Intervention | 13.26 ± 12.39<br>13.28 ± 11.97 | -----<br>----- | -----<br>----- | -----<br>----- | ---- | 15.49 ± 11.98<br>5.83 ± 6.98 | 2.31(0.11/4.38)<br>-7.45(-10.38/-4.52) | 0.039<br>0.001* | 0.18<br>0.76 |
| Actual H. VAS<br>Control<br>Intervention | 2.04 ± 1.76<br>1.75 ± 1.74 | 2.51 ± 2.62<br>0.74 ± 1.22 | 0.47(-0.39/1.34)<br>-1.01(-1.69/-0.32) | 0.544<br>0.002* | 0.21<br>0.67 | 1.82 ± 2.08<br>0.86 ± 1.54 | -0.23(-1.34/0.39)<br>0.88(-1.57/-0.18) | 0.999<br>0.009 | 0.11<br>0.54 |
| Usual H. VAS<br>Control<br>Intervention | 2.99 ± 1.46<br>3.42 ± 0.90 | -----<br>----- | -----<br>----- | -----<br>----- | ---- | 3.13 ± 1.86<br>1.65 ± 1.43 | 0.13(-0.30/0.57)<br>-1.76(-2.35/-1.17) | 0.540<br>0.001* | 0.07<br>1.48 |
| Worse H. VAS<br>Control<br>Intervention | 6.14 ± 1.71<br>6.27 ± 1.64 | -----<br>----- | -----<br>----- | -----<br>----- | ---- | 5.91 ± 2.11<br>4.46 ± 2.50 | -0.23(-0.98/0.52)<br>-1.81(-2.63/-0.98) | 0.542<br>0.001* | 0.12<br>0.85 |
| Better H. VAS<br>Control<br>Intervention | 0.54 ± 1.05<br>0.55 ± 0.91 | -----<br>----- | -----<br>----- | -----<br>----- | ---- | 0.49 ± 0.89<br>0.18 ± 0.48 | -0.04(-0.24/0.15)<br>-0.37(-0.65/-0.09) | 0.639<br>0.010 | 0.05<br>0.51 |
| Flexion<br>Control<br>Intervention | 51.35 ± 10.42<br>47.55 ± 14.12 | 45.64 ± 10.13<br>53.27 ± 11.62 | -5.71(-9.36/-2.07)<br>5.72(0.81/10.63) | 0.001*<br>0.017 | 0.56<br>0.44 | 46.77 ± 14.12<br>52.27 ± 12.00 | -4.59(-9.11/-0.067)<br>4.72(0.14/9.31) | 0.046<br>0.041 | 0.37<br>0.36 |
| Extension<br>Control<br>Intervention | 55.53 ± 11.61<br>50.60± 11.45 | 50.46 ± 11.64<br>59.32 ± 14.23 | -5.07(-9.59/-0.55)<br>8.72(2.10/13.32) | 0.023<br>0.001* | 0.44<br>0.67 | 49.53 ± 11.45<br>57.00 ± 12.73 | -6.00(-9.89/-2.10)<br>6.40(2.33/10.46) | 0.001*<br>0.001* | 0.52<br>0.53 |
| Side-Bending (R)<br>Control<br>Intervention | 32.69 ± 8.90<br>33.02± 8.34 | 31.51 ± 7.50<br>38.60 ± 8.47 | -1.17(-3.16/0.80)<br>5.57(2.80/8.36) | 0.432<br>0.001* | 0.14<br>0.66 | 29.53 ± 8.34<br>36.62 ± 9.53 | -3.15(-5.31/-0.93)<br>3.60(1.18/6.01) | 0.003*<br>0.002* | 0.36<br>0.40 |
| Side-Bending (L)<br>Control<br>Intervention | 34.76 ± 8.13<br>33.95 ± 7.85 | 33.94 ± 7.05<br>40.62 ± 8.83 | -0.82(-3.06/1.42)<br>6.67(4.45/8.89) | 0.999<br>0.001* | 0.11<br>0.80 | 34.00 ± 7.78<br>38.75 ± 8.29 | -0.77(-3.23/1.69)<br>4.80(2.92/6.67) | 0.999<br>0.001* | 0.10<br>0.56 |
| Rotation (R)<br>Control<br>Intervention | 57.53 ± 9.22<br>56.55 ± 7.75 | 56.53 ± 9.07<br>63.30 ± 7.75 | -1.00(-4.32/2.32)<br>6.75(3.32/10.18) | 0.999<br>0.001* | 0.11<br>0.87 | 55.25 ± 9.77<br>61.30 ± 7.43 | -2.28(-4.32/2.32)<br>4.75(1.53/7.96) | 0.297<br>0.002* | 0.24<br>0.63 |
| Rotation (L)<br>Control<br>Intervention | 59.69 ± 9.33<br>58.72 ± 8.48 | 57.05 ± 8.40<br>65.62 ± 9.21 | -2.64(-5.96/0.68)<br>6.90(3.20/10.59) | 0.162<br>0.001* | 0.28<br>0.78 | 56.69 ± 9.24<br>64.15 ± 8.86 | -3.00(-6.13/0.13)<br>5.42(1.83/9.02) | 0.065<br>0.002* | 0.32<br>0.62 |
| UCS Flexion<br>Control<br>Intervention | -0.84 ± 10.17<br>-2.20 ± 8.88 | -2.56 ± 10.95<br>3.72 ± 9.51 | -1.72(-3.75/0.32)<br>5.92(3.69/8.15) | 0.124<br>0.001* | 0.16<br>0.64 | -2.41 ± 10.12<br>1.10 ± 9.79 | -1.56(-3.75/0.32)<br>3.30(0.92/5.67) | 0.158<br>0.004* | 0.15<br>0.35 |
| UCS Extension<br>Control<br>Intervention | 35.92 ± 10.00<br>34.92 ± 6.79 | 35.12 ± 8.25<br>38.60 ± 6.19 | -0.26(-3.37/1.78)<br>3.67(1.64/5.70) | 0.999<br>0.001* | 0.09<br>0.56 | 35.89 ± 8.96<br>38.80 ± 6.31 | -0.79(-3.37/1.78)<br>3.87(1.68/6.07) | 0.999<br>0.001* | 0.01<br>0.59 |

Abbreviations: H: Headache, VAS: visual analogue scale, R: right, L: left, UCS: upper cervical spine, ES: effect size
<sup>a</sup> Comparison between baseline and after 3<sup>rd</sup> intervention. <sup>b</sup> Comparison between baseline and 1-month follow up. \* $p < 0.005$

The Table 3 show the differences between treatment groups in each outcome at T1 and T2 follow-ups.

|  | Follow up T1 |  |  | Follow up T2 |  |  |
| --- | --- | --- | --- | --- | --- | --- |
|  | Difference between Baseline<br>Mean (95% CI) | p<sup>a</sup> | ES | Difference Between Baseline<br>Mean (95% CI) | p<sup>b</sup> | ES |
| Frequency of headache | ----- | --- | --- | -9.66 (-14.04/-5.28) | 0.001* | 0.79 |
| Actual headache VAS | -1.48 (-2.36/-0.60) | 0.001* | 0.83 | -0.65 (-1.51/0.19) | 0.001* | 0.38 |
| Usual headache VAS | ----- | --- | --- | -1.89 (-2.62/-1.16) | 0.001* | 1.56 |
| Worse headache VAS | ----- | --- | --- | -1.57 (-2.67/-0.48) | 0.005* | 0.93 |
| Better headache VAS | ----- | --- | --- | -0.32 (-0.66/0.01) | 0.056 | 0.32 |
| Flexion | 11.44 (6.55/16.32) | 0.001* | 0.91 | 9.31 (4.19/14.43) | 0.001* | 0.74 |
| Extension | 13.8 (8.67/18.93) | 0.001* | 1.18 | 12.4 (7.91/16.88) | 0.001* | 1.06 |
| Side-Bending (R) | 6.75 (4.02/9.48) | 0.001* | 0.77 | 6.75 (4.14/9.36) | 0.001* | 0.77 |
| Side-Bending (L) | 7.49 (4.98/10.00) | 0.001* | 0.93 | 5.57 (3.11/8.02) | 0.001* | 0.69 |
| Rotation (R) | 7.75 (3.94/11.55) | 0.001* | 0.90 | 7.03 (3.32/10.74) | 0.001* | 0.82 |
| Rotation (L) | 9.54 (5.58/13.50) | 0.001* | 1.06 | 8.42 (4.62/12.22) | 0.001* | 0.92 |
| UCS Flexion | 7.64 (5.23/10.04) | 0.001* | 0.79 | 4.86 (2.41/7.31) | 0.001* | 0.50 |
| UCS Extension | 4.46 (1.86/7.07) | 0.001* | 0.52 | 3.90 (1.40/6.39) | 0.003* | 0.45 |

Abbreviations: VAS: visual analogue scale, R: right, L: left, UCS: upper cervical spine, ES: effect size
<sup>a</sup> Comparison between baseline and 48h follow up. <sup>b</sup> Comparison between baseline and 1-month follow up. \*p<0.005

## Discussion

This randomized controlled trial aimed to evaluate the effectiveness of three treatment sessions of diacutaneous fibrolysis in headache frequency, pain intensity, cervical pressure pain thresholds and cervical mobility in patients with tension-type headache.

The subjects who received the intervention obtained an improvement in the frequency of headache episodes, which is more remarkable considering that the control group worsened compared to the episodes suffered in the same period.

An improvement in the intensity of the headache was obtained for the four recorded situations, this reduction being in favor of the intervention group. Furthermore, cervical range of motion improved and the sensibility of the assessed pericranial tissue decreased.

In a recent review and meta-analysis, Kamonseki et al. [13] have found that other techniques of manual therapy such as soft tissue treatment and dry needling have been shown to be superior to no treatment on frequency of pain in tension-type headache. Soft tissue treatment techniques have also been shown to be more beneficial than other types of treatment techniques, but with less difference.

The intervention group’s frequency decreased significantly (p <0.001) in 7.45 days, while the control group showed an increase of 2.31 days. It seems that the treatment of the musculature applied in the cranio-cervical region is an effective strategy to reduce the frequency of headache in subjects with tension-type headache. Other authors collect the frequency of headache suffered in two weeks or consider a specific percentage reduction in episodes to classify them as having improved [28]. Heterogeneity in the collection of frequency makes it difficult to compare results between studies.

After applying the intervention protocol, a decrease in the usual headache, and the worst headache felt in the last 4 weeks was observed, the decrease was 1.7 for the usual headache and 1.8 for the worst headache. Manual therapy has been suggested to be effective in reducing headache intensity in this type of subjects. Different authors have obtained changes of 1 to 3.1 cm in this scale with manual therapy treatment. These studies were carried out between eight and twenty treatment sessions, compared to the three applied in our study [29–31]. The study carried out by Toro Velasco et al. [32] would be the treatment that most closely resembles ours in terms of the dose and treatment goal.

Two suboccipital massage sessions were performed, obtaining an immediate improvement of 1 cm in the visual analogue scale. Our treatment achieved improvements superior to this but with a slightly higher dose.

Pericranial tenderness is one of the common symptoms in patients with tension-type headache. The pressure pain threshold is one of the most common ways of objectifying pain in mechanosensitive points. In the review published by Fernández de Las Peñas et al. [5], it is observed that variables as the gender or the type of tension-type can affect the pressure pain thresholds, sensitivity to pressure pain was widespread only in chronic tension-type headache and women had lower pressure than men. Ferragut Garcías et al. [21] carried out a protocol of soft-tissue techniques and neural mobilization evaluating the effect on the pressure pain thresholds on two temporal points and the supraorbital region. After six treatment sessions, the neural mobilization group and the soft-tissue group obtained improvements in pressure pain thresholds, achieving an increase of 41.7% to 67.5% in the pressure pain thresholds’ values compared with the baseline. In this study, an increase in mechanosensitivity to pressure was observed in the control group for all the points assessed both between the baseline and T1 and between the baseline and T2.

On the other hand, we observed a decrease in mechanosensitivity for all the points evaluated in the intervention group except for the trapeziometacarpal joint between baseline and T1. As points to be highlighted, it is observed that in the intervention group between the baseline and T2, the mechanosensitivity to pressure had increased by 15.7% for the left temporal muscle, by 19.95% in the right frontal muscle, by 20.78% in the left frontal muscle and by 17.34% on the right parietal. For these points, a change greater than 15% was obtained concerning baseline for temporal muscle, frontal muscle, and right parietal suture. This percentage of change is considered the minimum clinically relevant change for pressure pain thresholds [33]. After local treatment, this widespread hypoalgesic response has been suggested to be produced by activating supraspinal pain inhibitory mechanisms, which are activated by mechanical force from manual therapy interventions [34].

It should be noted that for all movements, the intervention group improved range of motion. However, the control group maintains the range of cervical motion or even in some cases, experiences a reduction. Despite these trends, in most cases the minimum change detectable value is not reached, calculated with the CROM tool for subjects with cervical pain due to Fletcher et al. [35].

Considering these promising findings, it could be recommended the use of diacutaneous fibrolysis treatment in tension-type headache. It seems that the three treatments with this type of therapy reduces the headache pain intensity after the intervention protocol and is maintained at one-month follow-up (T2). It also improves the cervical and upper cervical range of motion in all cervical movements. The pressure pain thresholds increase therefore, it seems that the local and distal hyperalgesia decrease.

The present study has some limitations that need to be addressed. Our sample included subjects with medical diagnosis of tension-type headache without differentiating between episodic or chronic type. It is possible that the differentiation between these subclassifications may be important in terms of the improvement of each type of patient [10].

Another limitation of this study has been the impossibility to blind the physiotherapist with respect to the applied interventions.

There is another limitation regarding the follow-up, which may have been brief if considering that some subjects suffered from chronic tension-type headache.

For future studies, it would be advisable to extend the follow-up period to analyze the long-term effects that treatment with diacutaneous fibrolysis may have, as well as to associate it with soft tissue self-treatment techniques that can maximize the effect obtained.

## Conclusions

Diacutaneous fibrolysis is effective to improve short-term and one month follow-up headache frequency and intensity and provides improvements in range of motion in patients with tension-type headache.

The application of three sessions of diacutaneous fibrolysis in the cervico-dorsal and the orofacial areas in patients with tension-type headache produced a self-perceived improvement compared to control subjects.

## Data Availability

All relevant data are within the manuscript and its Supporting Information files.

## Acknowledgments

Thanks to all the patients who participated in this study.

